# Comparative Assessment of the Prevalence, Practices and Factors Associated with Self-medication with Antibiotics in Africa: A Systematic Review and Meta-analysis

**DOI:** 10.1101/2021.03.24.21254283

**Authors:** Eugene Vernyuy Yeika, Brecht Ingelbeen, Ben-Lawrence Kemah, Frankline Sevidzem Wirsiy, Joseph Nkeangu Fomengia, Marianne van der Sande

## Abstract

**Background:** Self-medication with antibiotics (SMA) is a common practise in many parts of Africa, but its magnitude and drivers are not well-known. This systematic review evaluated and compared the prevalence, reasons, sources, and factors associated with of SMA within African sub-regions.

**Methods:** An electronic search of PubMed and Google Scholar databases was performed for observational studies conducted between January 2005 and February 2020. Two reviewers independently screened the abstracts and full texts using the PRISMA flowchart and equally performed quality assessment. Both quantitative and qualitative syntheses of selected studies were carried out.

**Results:** A total of 40 studies from 19 countries were selected for qualitative synthesis. The prevalence of SMA in Africa ranged from 12.1% to 93.9% with a median prevalence of 55.7% (IQR 41%-75%). Western Africa was the sub-region with the highest prevalence 70.1% (IQR 48.3%-82.1%), followed by Northern Africa with 48.1% (IQR 41.1-64.3%). We identified 27 different antibiotics used for self-medication from 13 different antibiotic classes. Penicillins were the most frequently used antibiotics followed by Tetracyclines and Fluoroquinolones. The most frequent indications for SMA were upper respiratory tract symptoms/infections (27 studies). Common sources of antibiotics used for self-medication were community pharmacies (31 studies), family/friends (20 studies), leftover antibiotics from previous treatments (19 studies), and patent medicine stores (18 studies). Commonly reported factors associated with SMA were no education/ low educational status (9 studies), male gender (5 studies), and low income / unemployment (2 studies).

**Conclusions:** The prevalence of SMA in Africa is high and varies across sub-regions with Western Africa having the highest prevalence. Drivers of SMA are complex comprising of socio-economic factors, limited access to healthcare coupled with absence or poorly implemented policies regulating antibiotic sales.

## BACKGROUND

Self-medication with antibiotics (SMA) is the use of antibiotics to treat self-recognized disorders without prior consultation by a qualified health professional (1). SMA can lead to inappropriate antibiotic use and other problems such as masking of underlying symptoms, delaying or misleading diagnoses, drug interactions and accelerating the emergence and spreading of antimicrobial resistance (2–4). The World Health Organization defines inappropriate antibiotic use as using antibiotics without proper indication, or administering wrong dosages, incorrect treatment duration, late or absence of downscaling of treatment, poor adherence to treatment, and use of poor quality or substandard antibiotics (5). In Low and Middle-Income Countries (LMIC), it is estimated that about 80% of antibiotics are used in communities, outside of official health care facilities, of which about 20-50% are used inappropriately (6). Other issues related to SMA are the wastage of economic resources from prolonged treatment durations due to incorrect management of infections and adverse reactions (1). Africa has been hardest hit by the growing antimicrobial resistance pandemic (7,8). This region carries a high burden of infectious diseases which compounds the growing weight of non-prescription sales and inappropriate use of antibiotics, and thus also its aforementioned challenges notably accelerating antimicrobial resistance (9). It was estimated in 2011 that more than half of the antibiotics used in communities especially in Africa were sold without a prescription (10). This review aimed to evaluate the magnitude and drivers of SMA amongst different populations in Africa and to generate evidence for recommendations to control and reduce inappropriate use of antibiotics.

## METHODS

### Search strategy

An electronic systematic search of the Medline through PubMed and Google Scholar databases was performed following the PRISMA statement (11). Search terms or keywords were identified through a pilot literature search and Boolean operators were used to combine these terms to come up with a search strategy [Table 1]. Medical Subject Headings (MeSH) were used to synchronizes synonymous terms in PubMed. We excluded reviews, animal models, editorials, letters, opinions or comment publication types. To ensure that no similar review had been registered or previously carried out, a preliminary scoping search was done on the following registries: International Prospective Register of Systematic Reviews (PROSPERO), International Platform of Registered Systematic Review and Meta-Analysis Protocols (INPLASY), Research Registry and Cochrane Library of Systematic Reviews and also on the PubMed and Google Scholar databases.

**Table 1:** Search strategy

| Database | Search mode | Search term syntax |
| --- | --- | --- |
| Medline via PubMed | All fields | ((("Anti-Bacterial Agents"[Mesh]) AND "Self Medication"[Mesh]) AND "Nonprescription Drugs"[Mesh]) AND "Drug Misuse"[Mesh]) AND "Africa"[Mesh] |
| Google Scholar | Articles | (antimicrobial* OR antibacterial* OR antibiotic*) AND ("self medication" OR self-medication OR "non prescription" OR non-prescription OR "over-the-counter OR "inappropriate") AND (determinants OR "associated factors") -Asia -Europe - America -Review |

### Selection of articles for review

Articles included in this review were selected using the PRISMA Flow Diagram (12). Two reviewers EVY and JNF, independently screened studies against the eligibility criteria and discrepancies were resolved through a third reviewer BI. Titles and abstracts of all records identified through database searches were screened for duplicates and studies that met the inclusion criteria were selected. These included observational studies carried out on SMA in Africa between January 2005 and February 2020. Studies on SMA conducted in other regions, dissertations on SMA, studies on general SM, studies on aggregated non-prescription antibiotic sales data, and studies with full-text published in languages different from English or French were excluded. Additional articles that met the inclusion criteria were identified by reading-through reference lists of studies eligible for full-text review. The full-text of selected studies were reviewed and studies not relevant to research questions, qualitative studies, and studies of which the full-text could not be retrieved were excluded.

### Assessment of the quality of included studies

Quality appraisal of studies selected for full-text review was done using the risk of bias in prevalence studies evaluation tool by Hoy et al, appraising the studies on 9 criteria (13).

### Data Extraction

Titles and abstracts of studies selected retrieved were saved in Mendeley. The following characteristics were extracted using a spreadsheet in excel: the country, corresponding author, year of publication, study site, study design, sampling strategy, recall period, sample size and response rate. The prevalence of SMA, type of antibiotics used for self-medication, reasons for non-SMA, sources of antibiotics, and factors associated with SMA were also extracted.

### Data syntheses

Both qualitative and quantitative syntheses were performed.

In the qualitative synthesis, we generated descriptive summaries of all studies and outcomes of interest. The WHO AWaRe Classification (14) was used to group reported antibiotics. Reasons for SMA were analyzed using the modified conceptual framework of access to healthcare (15). Prevalence estimates were summarized using medians and interquartile ranges, and box-whiskers plots were generated to compare their differences.

The quantitative synthesis was conducted only on studies carried out within household using the ‘metafor’ package of R software (version 3.6.1). Random-effect model was used to calculate the weight of each study and Freeman-Tukey double arcsine transformation was use to stabilise the variance in the proportions of individual studies. Heterogeneity was checked by Cochran’s Q-test and quantified by the I^2^. Heterogeneity was considered present and statistically significant when I^2^>50% and P-value<0.05. Findings were displayed graphically using Forest Plots. To verify publication bias, funnel Plots were constructed using Double Arcsine transformed proportions.

## RESULTS

### Search results

The database search was done on the 26^th^ February, 2020 and a total of 6940 citations were identified; 240 citations through PubMed and 6700 citations through Google Scholar. After adjusting for duplicates, 212 citations were discarded. The titles and abstracts of the remaining 4,218 studies were screened and 4,174 records were disqualified as they did not meet the inclusion criteria. Reference mining was carried out on the selected 44 studies and 8 additional studies were identified making a total of 52 studies selected for full-text review. After reviewing the full-text of the selected studies, 12 studies were further excluded. The remaining 40 selected studies served in qualitative synthesis. The 15 studies that were carried out in households, were selected for quantitative synthesis [Figure 1].

**Figure 1.**
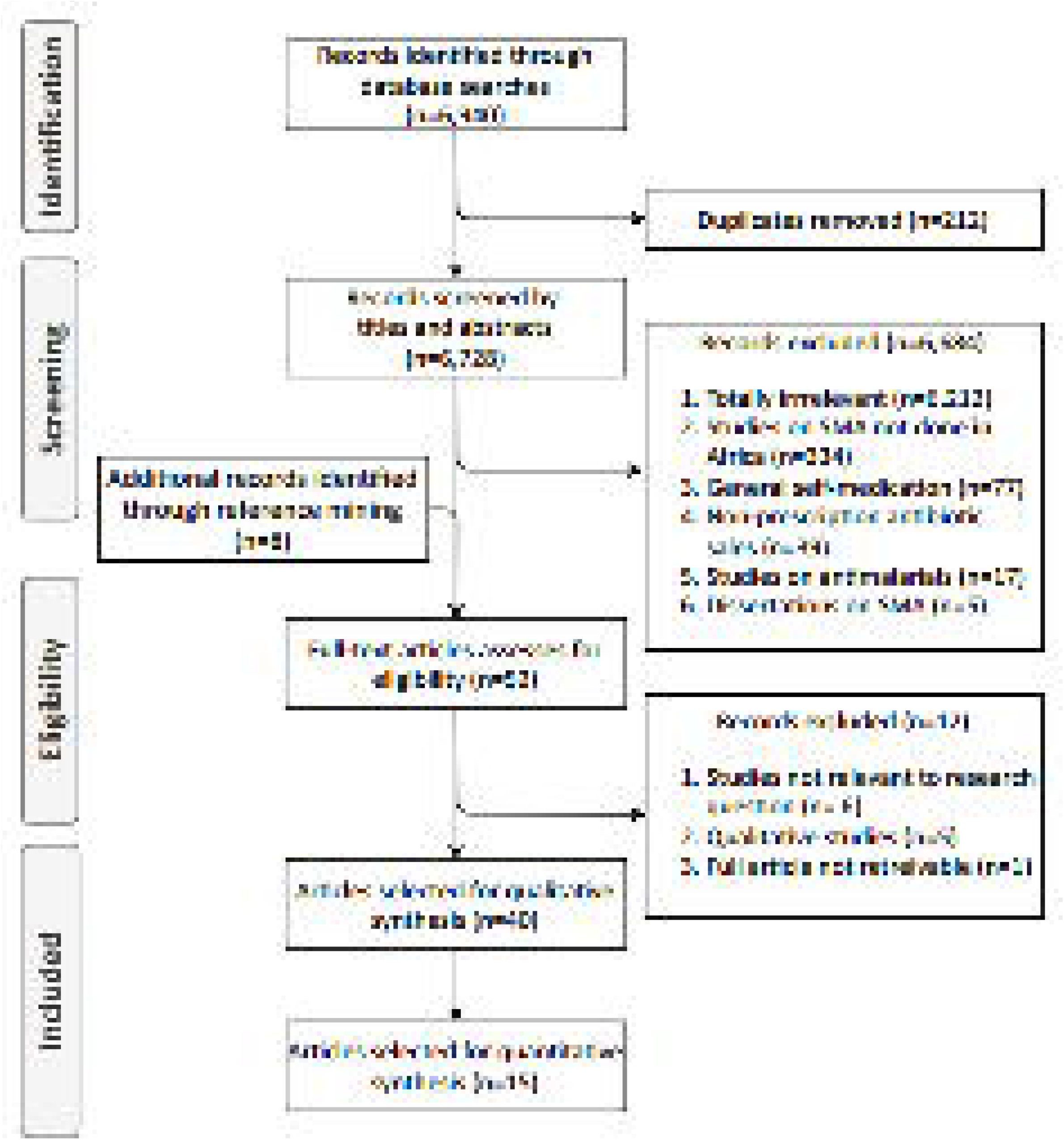

### Study characteristics

The 40 studies included in this review were from 19 African countries and all the 5 sub-regions: 7 from Northern Africa (16–22), 3 from Central Africa (23–25), 14 from Western Africa (26–39), 15 from Eastern Africa (40–53) and 1 from Southern Africa (54) [Figure 2]. Thirty-seven of these studies were cross-sectional surveys and 3 mixed methods (i.e. cross-sectional surveys with qualitative work) (31,44,49). Fifteen of these studies were carried out in households, 13 studies academic settings (universities), 4 studies in pharmacies, 3 studies in health facilities, and 5 studies in other settings (markets, streets, shopping malls, offices). All studies together included 21,358 participants with sample sizes ranging from 110 - 1750. The recall period used in data collection ranged from 3 days to 10 months and was reported by 32 studies [Table 2].

**Figure 2.**
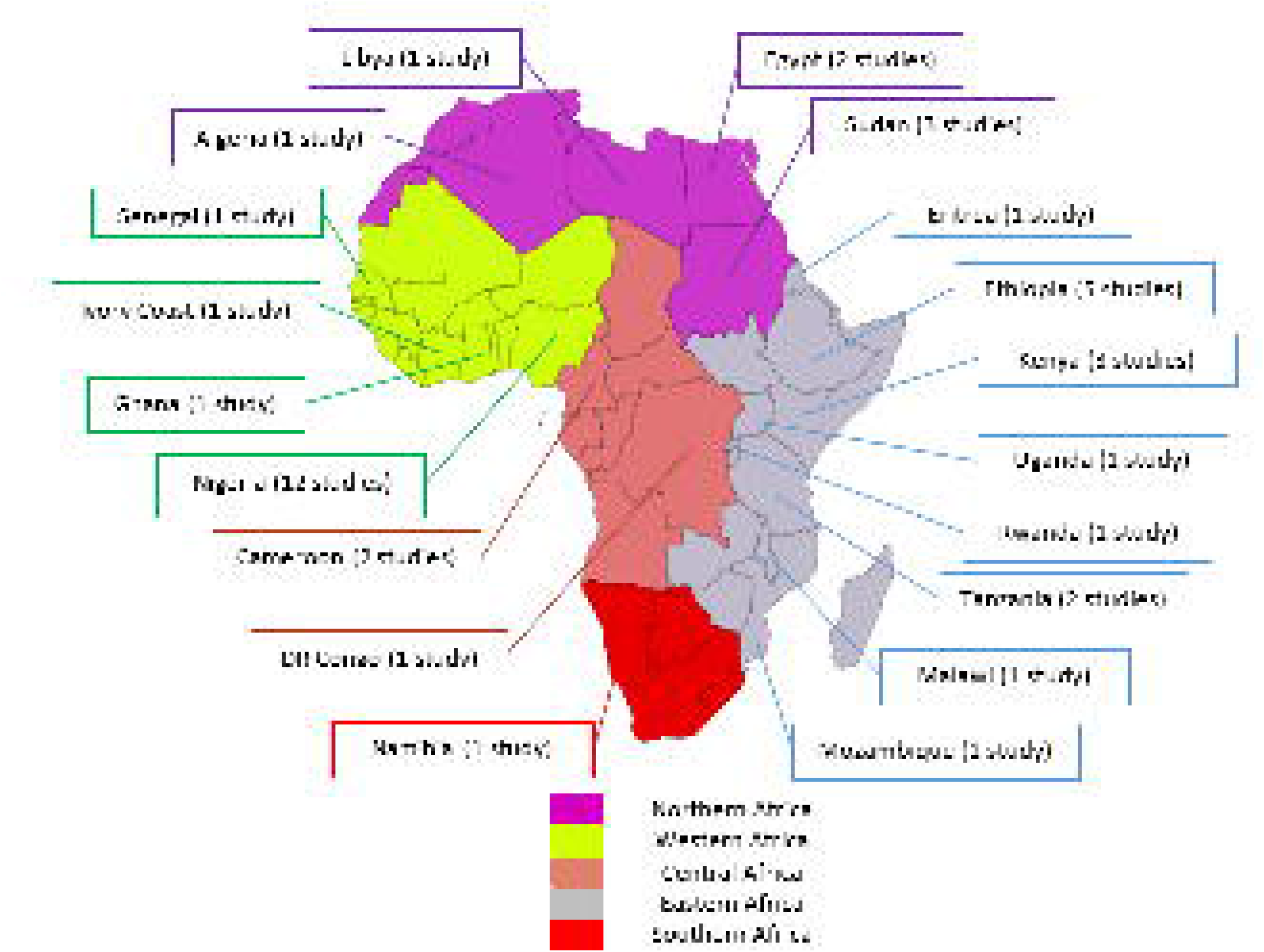

**Table 2:** Study characteristics and prevalence rates of SMA

| Reference | Country | Study site | Study participants | Sampling method | Sample size | Recall period/ weeks | Response rate | Prevalence | 95% CI |
| --- | --- | --- | --- | --- | --- | --- | --- | --- | --- |
| *Gacem et al, 2015* (16) | Algeria | Pharmacies | General public | Simple random | 159 | 2 | NR | 12.6% | 07.4 - 17.8 |
| *Elden et al, 2020* (17) | Egypt | Universities | Students | Multistage | 600 | 40 | 94% | 77.7% | 74.4 - 81.0 |
| *El-Hawy et al, 2017* (18) | Egypt | Clubs, cafes, streets | General public | Convenient | 400 | 8 | 89.7% | 64.3% | 59.6 - 69.0 |
| *Ghaieth et al, 2015* (19) | Libya | Universities | Students | NR | 665 | 16 | 55% | 46% | 42.2 - 49.8 |
| *Awad et al, 2005* (20) | Sudan | Households | General public | Multistage stratified cluster, simple random | 1750 | NR | 89.7% | 48.1% | 45.8 - 50.4 |
| *Awad et al, 2007* (21) | Sudan | Universities | Undergraduate students | Multistage stratified cluster, systematic random | 1121 | 24 | 86.2% | 55% | 52.1 - 57.9 |
| *Ahmed et al, 2014* (22) | Sudan | Households | General public | Simple random | 442 | 36 | NR | 41% | 36.4 - 45.6 |
| *Ngu et al, 2018* (23) | Cameroon | Hospitals | Patients with respiratory tract infection | Convenient | 308 | 24 | NR | 41.9% | 36.4 - 47.4 |
| *Amin et al, 2019* (25) | Cameroon | Health facilities | Patients | NR | 329 | 16 | NR | 68.4% | 63.4 - 73.4 |
| *Bunduki et al, 2017* (24) | DR Congo | Universities | Students | Convenient | 500 | 3 | 86% | 90.7% | 88.2 - 93.2 |
| *Donkor et al, 2012* (26) | Ghana | Universities | Students | Stratified sampling, Convenient | 600 | 32 | 90% | 70% | 66.3 - 73.7 |
| *Olayemi et al, 2010* (27) | Nigeria | Universities | Undergraduate students | Simple random | 430 | NR | 65.8% | 56.9% | 52.2 - 61.6 |
| *Abdulraheem et al, 2016 (32)* | Nigeria | Health Centres | All patients | Simple random | 1150 | 24 | 93.9% | 82.2% | 80.0 - 84.4 |
| *Badger-Emeka et al, 2018 (33)* | Nigeria | Households | General public | Convenient | 400 | 28 | NR | 86% | 83.2 - 89.8 |
| *Ehigiator et al, 2010 (34)* | Nigeria | Universities | Dental students | NR | 208 | 12 | 96.2% | 53.5% | 46.5 - 60.3 |
| *Israel et al, 2015 (35)* | Nigeria | Ministries, departments, units | Civil servants | Simple random | 526 | NR | 89.5% | 93.9% | 91.9 - 95.9 |
| *Fadare et al, 2011 (36)* | Nigeria | Universities | Medical students | Convenient | 183 | 4 | 83.2% | 38.8% | 31.7 - 45.9 |
| *Sapkota et al, 2010 (37)* | Nigeria | Universities | Students | Three-stage cluster, simple random | 740 | NR | 95.4% | 24% | 20.9 - 27.1 |
| *Umar et al, 2018 (38)* | Nigeria | Universities | Paramedical students | Stratified | 115 | NR | 82% | 81.9% | 74.9 - 88.9 |
| *Yusuf et al, 2019 (39)* | Nigeria | Households | General public | Simple random | 300 | 8 | 85.3% | 70.3% | 65.1 - 75.5 |
| *Ajibola et al, 2018 (28)* | Nigeria | Hall of residence | Community residents & undergraduate students | Convenient | 1450 | 8 | 84.8% | 43% | 40.5 - 45.5 |
| *Khalid et al, 2019 (29)* | Nigeria | Universities | Pharmacy students | Purposive | 217 | 4 | 100% | 92.2% | 88.6 - 95.8 |
| *Bassoum et al, 2019 (30)* | Senegal | Bus station | General public | Convenient | 400 | 4 | 100% | 75% | 70.8 - 79.2 |
| *Hounsa et al, 2010 (31)* | Ivory Coast | Pharmacies | General public | Simple random | 1123 | 24 | NR | 59.7% | 56.8 - 62.6 |
| *Ateshim et al, 2019 (40)* | Eritrea | Households | General public | Two-stage cluster, systematic random | 580 | 12 | 99.5% | 45.1% | 41.1 - 49.1 |
| *Bogale et al, 2019 (55)* | Ethiopia | Households | General public | Multistage, systematic random | 605 | 12 | 98.3% | 67.3% | 63.6 - 71.0 |
| *Erku et al, 2017*<br>(41) | Ethiopia | Households | General public | Multistage,<br>stratified random,<br>systematic random | 720 | 8 | 90.3% | 63.5% | 60.0 – 67.0 |
| *Eticha et al, 2014*<br>(46) | Ethiopia | Universities | Undergraduate<br>Students | Stratified, simple<br>random | 422 | 4 | 96.4% | 44.5% | 39.8 - 49.2 |
| *Nyambega et al,*<br>*2017*(47) | Kenya | Markets,<br>shopping<br>malls and<br>households | General public | Simple random | 385 | 8 | 78% | 60% | 55.1 - 64.9 |
| *Owour et al, 2015*<br>(48) | Kenya | Households | General public | Cluster, systematic<br>random | 350 | 3/7 | NR | 76.9% | 72.5 - 81.3 |
| *Sambakunsi et al,*<br>*2019* (49) | Malawi | Households | General public | Weighted cluster<br>random,<br>snowballing | 110 | NR | 95.5% | 41% | 31.8 - 50.2 |
| *Tuyishimire et al,*<br>*2019*(50) | Rwanda | Universities | Undergraduate<br>students | Simple random | 570 | NR | NR | 12.1% | 09.4 - 14.8 |
| *Horumpende et al,*<br>*2018* (51) | Tanzania | Households | General public | Systematic random | 300 | 4 | NR | 58% | 52.4 - 63.6 |
| *Kajeguka et al,*<br>*2017*(52) | Tanzania | Households | General public | Simple random | 300 | 12 | NR | 55.7% | 50.1 - 61.3 |
| *Ocan et al, 2014*<br>(53) | Uganda | Households | General public | Multistage cluster,<br>Simple random | 892 | 8 | 99.1% | 75.7% | 72.9 - 78.5 |
| *Gebeyehu et al,*<br>*2015*(42) | Ethiopia | Households | General public | Systematic random | 1082 | 8 | 98.3% | 18% | 15.7 - 20.3 |
| *Gebrekiros et al,*<br>*2017* (43) | Ethiopia | Drug retail<br>outlets | General public | Stratified, simple<br>random | 829 | 8 | 94% | 47.1% | 43.7 - 50.5 |
| *Owuor et al, 2019*<br>(44) | Kenya | Households | General public | Two-stage cluster,<br>systematic random | 380 | NR | 83.2% | 20.9% | 16.8 – 25.0 |
| *Mate et al, 2019*<br>(45) | Mozambique | Households | General public | Three-stage<br>cluster, Random | 1091 | 12 | 73.1% | 20.9% | 18.5 – 23.3 |
| *Pereko et al, 2015*<br>(54) | Namibia | Pharmacies | General public | Simple random | 600 | 16 | 74.3% | 15.47% | 12.6 - 18.4 |

NR = Not Reported, CI = Confidence Interval

### Quality assessment of included studies

Forty studies were assessed for risk of bias. Three of these studies met all the 9 quality criteria in the assessment tool. Twenty-four studies showed a low-level risk of bias, 15 studies showed a moderate-level risk of bias, and 1 study presented with a high-level risk of bias.

### Prevalence of SMA

The prevalence of SMA ranged from 12.1% to 93.9%. Twenty-three studies reported prevalence estimates above 50%, 13 studies above 70% and 3 studies above 90% (90.3% in the Democratic Republic of Congo, 93.9% and 92.2% in Nigeria). Prevalence estimates of less than 20% SMA were reported in 4 studies. The overall median prevalence was 55.7% (IQR: 41%, 75%) and the median prevalence for Northern Africa was 48.1% (IQR: 41.1, 64.3%), Western Africa was 70.1% (IQR: 48.3%, 82.1%) and Eastern Africa was 47.1% (IQR: 31%, 65.4%). The prevalence of studies conducted in households ranged from 18% - 86% with a median prevalence of 48.1% (IQR: 41%, 73%). A meta-analysis of these studies revealed a pooled prevalence estimate of 51.5% (95% CI: 40.1%, 62.8%). The I^2^ = 99.1% (p<0.0001) indicative of pronounced heterogeneity. This means that the variation across studies is higher than that observed by chance, hence the pooled proportion of SMA was incongruous. The summary of results is presented in the forest plot [Figure 3].

**Figure 3.**
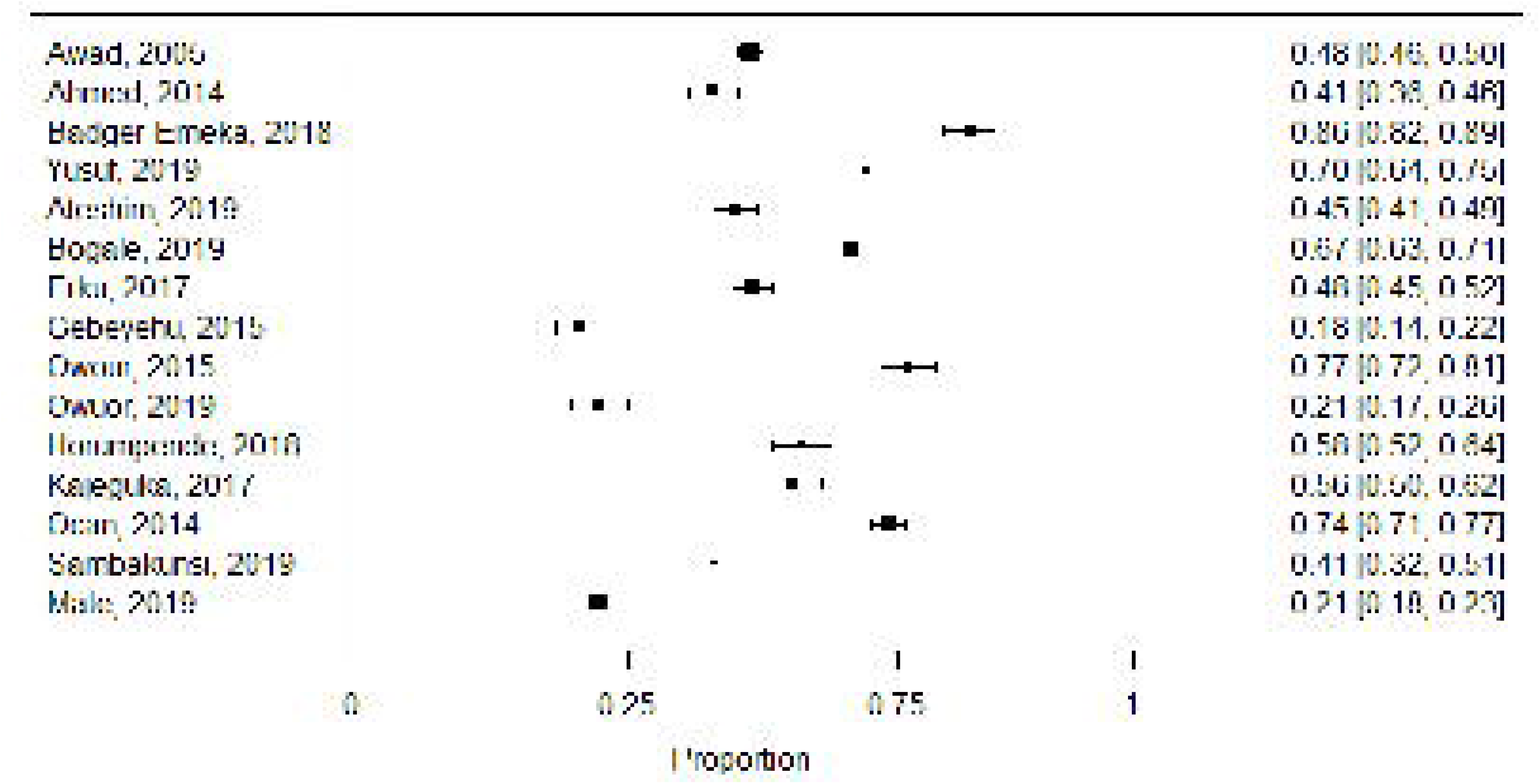

We analyzed the pooled estimates of the sub-regions exploring the cause of the observed heterogeneity. The pooled prevalence estimate for Northern Africa was 44.5% (95% CI: 18.3%, 72.5%), Western Africa was 78.5% (95% CI: 51.4%, 96.4%), and Eastern Africa was 47.5% (95% CI: 35.4%, 59.8%). High residual heterogeneity was equally observed with I^2^ = 98.8% (p<0.0001) indicating that the observed heterogeneity was not due to sub-regions. Funnel plots showed an asymmetric distribution of studies with most of them falling out of the funnel indicative of publication bias [Figure 4].

**Figure 4.**
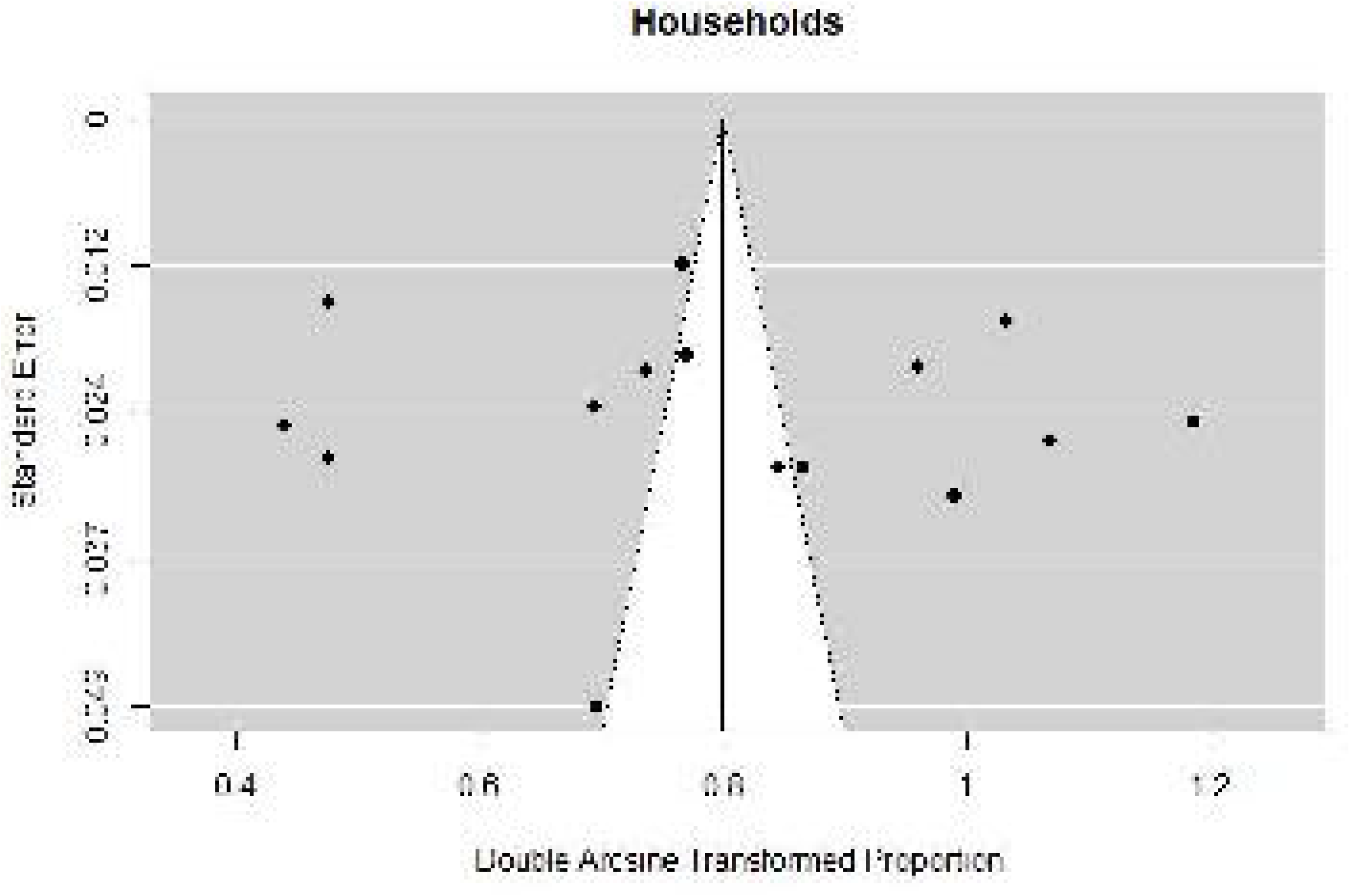

### Common antibiotics used in self-medication

Twenty-seven antibiotics from 13 different classes were identified used in SMA, and reported by 31 studies. The majority of these antibiotics (48%) belonged to the Access Group, 41% belonged to the Watch Group, and only one antibiotic belonged to the Reserve Group [Table 3]. The most frequently used classes of antibiotics were Penicillins (31 studies), Tetracyclines (25 studies), Fluoroquinolones (23 studies), Imidazoles (19 studies), Macrolides (10 studies), Amphenicols (9 studies) and Trimethoprim/Sulphonamides (17 studies) [Table 3].

**Table 3:** AWaRe classification of antibiotics used for SM

| **Class of antibiotic** | **Antibiotic used in SMA** | **AWaRe Group** |
| --- | --- | --- |
| **Penicillins**<br>(31 studies) | Amoxicillin | Access |
|  | Cloxacillin | Access |
|  | Flucloxacillin | Access |
|  | Ampicillin | Access |
|  | Penicillin | Access |
|  | Ampicillin/Cloxacillin | Not recommended |
|  | Amoxicillin/ clavulanic acid | Access |
| **Fluoroquinolones**<br>(23 studies) | Ciprofloxacin | Watch |
|  | Levofloxacin | Watch |
|  | Ofloxacin | Watch |
| **Trimethoprim/Sulphonamides** (17 studies) | Cotrimoxazole | Access |
| **Tetracyclines**<br>(25 studies) | Tetracycline | Access |
|  | Doxycycline | Access |
| **Macrolides**<br>(10 studies) | Erythromycin | Watch |
|  | Azithromycin | Watch |
| **Imidazoles**<br>(19 studies) | Metronidazole | Access |
| **Amphenicols**<br>(9 studies) | Chloramphenicol | Access |
| **Cephalosporins**<br>(4 studies) | Cefuroxime | Watch |
|  | Ceftriaxone | Watch |
|  | Cefixime | Watch |
| **Aminoglycosides**<br>(3 studies) | Gentamycin | Access |
|  | Streptomycin | Watch |
| **Aminocyclitols**<br>(1 study) | Spectinomycin | Access |
| **Nitrofurans**<br>(1 study) | Nitrofurantoin | Access |
| **Glycopeptides**<br>(1 study) | Vancomycin | Watch |
| **Polymyxins**<br>(1 study) | Polymyxin B | Reserve |

SMA: Self-medication with antibiotics

### Source of antibiotics used for self-medication

Thirty-two studies provided information on the main sources of antibiotics used for SMA. These include CPs (31 studies), family/friends (20 studies), leftover antibiotics from previous treatments (19 studies), PMS (18 studies), hospital pharmacies (8 studies), street vendors (7 studies), private health facilities (6 studies), healthcare workers (3 studies) and home medicine cabinets (2 studies) [Figure 5].

**Figure 5.**
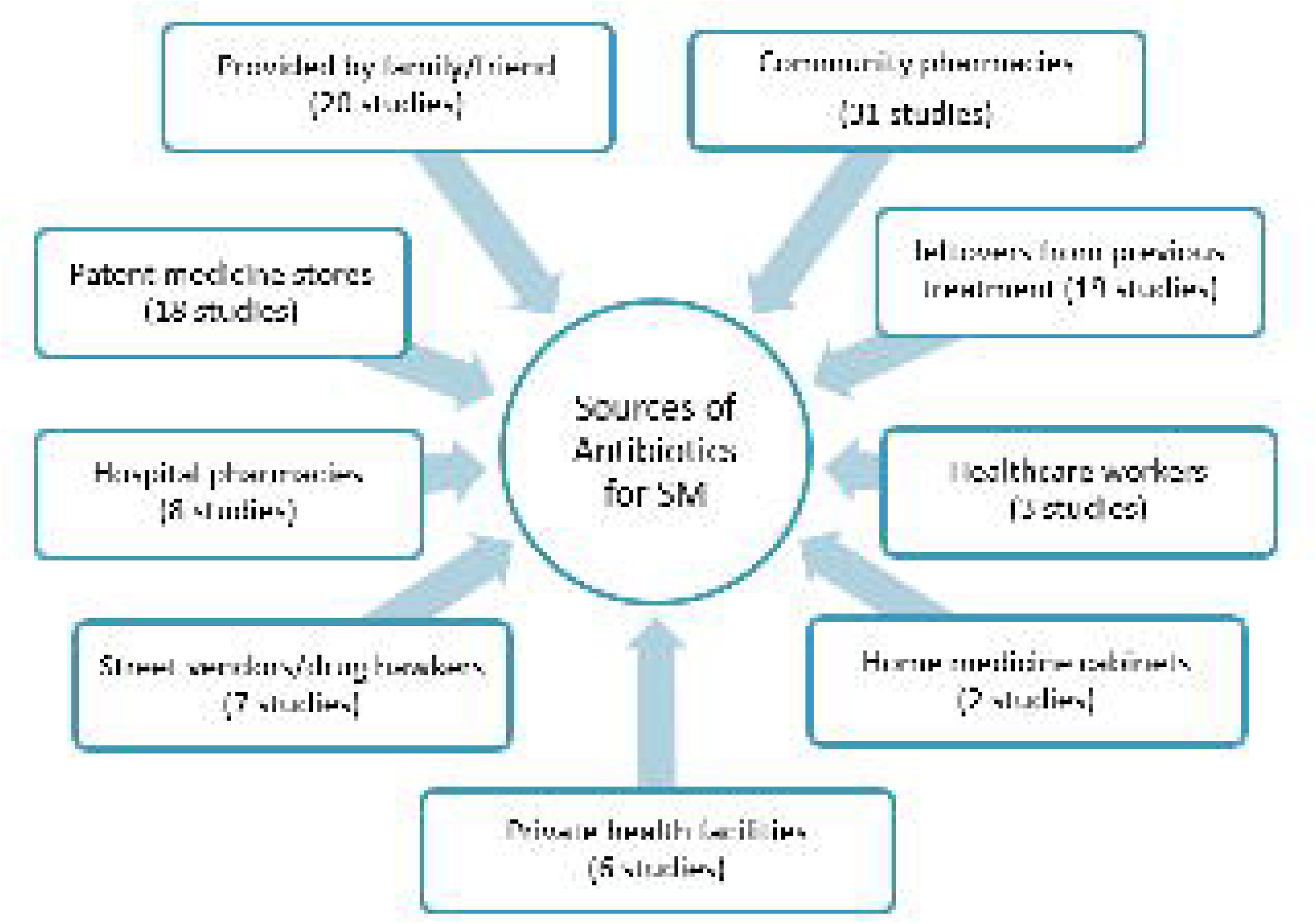

### Reasons for SMA

Twenty-nine studies reported reasons why people self-medicate with antibiotics. These reasons include past or prior experience with using similar symptoms or antibiotics (22 studies), additional cost incurred from facility charges (18 studies), long waiting time required to consult at health facilities (18 studies), illness perceived as mild by the patient (14 studies), advice from friend or relative (9 studies), lack of time to consult (8 studies), assumed knowledge on antibiotics use (5 studies), financial constraint (5 studies), nonchalant attitude of health workers (8 studies), lack of confidence in the healthcare system (6 studies), difficulty access to health facility due to remoteness (9 studies), easy access to antibiotics due to non-prescription sales (8 studies), emergency relief of symptoms (8 studies) and poorly staffed and equipped hospitals (3 studies) (Figure 6].

**Figure 6.**
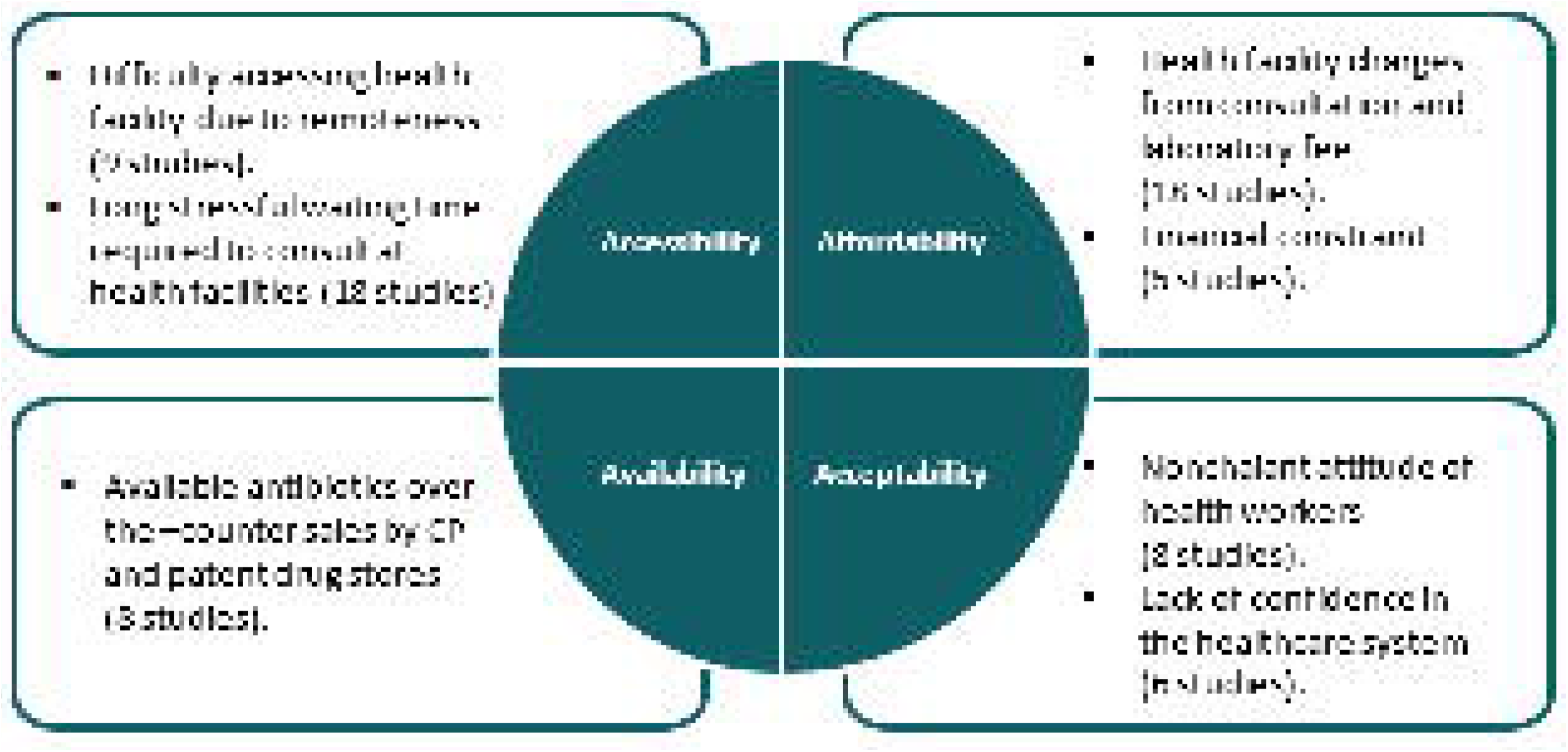

### Common symptoms/illnesses that resulted in SMA

The most common indications for SMA reported were upper respiratory tract symptoms/infections (common cold, cough, catarrh/runny nose, nasal congestion, sore throat, rhinitis, throat pain, tonsillitis) in 27 studies, followed by gastrointestinal tract symptoms (diarrhea, abdominal pain, vomiting) in 25 studies, fever or febrile illnesses in 18 studies, body aches (headache, toothache, joint pains, malaise) in 15 studies, skin injuries, infections and rashes/acne in 15 studies, urogenital tract symptoms in 10 studies, sexually transmitted infections in 5 studies, eye infections in 5 studies, dental infections in 4 studies, and menstrual symptoms in 3 studies.

### Factors associated with SMA

Twenty-one studies reported results of multivariable logistic regression analysis to determine factors associated with SMA. No education or low educational status were the most frequently reported factors by 9 studies (30,31,37,40–42,45,51,55). Five studies reported male gender (32,40,43,45,53), while 2 studies presented the female gender (20,51). Other associated factors reported were low income or unemployment (52,55), young age <30 years (17,42,55), middle-age 30 to 60 years (20,51), the remoteness of health facility (48,53), and perceived long waiting time at health facilities (43,53) [Table 4].

**Table 4:**
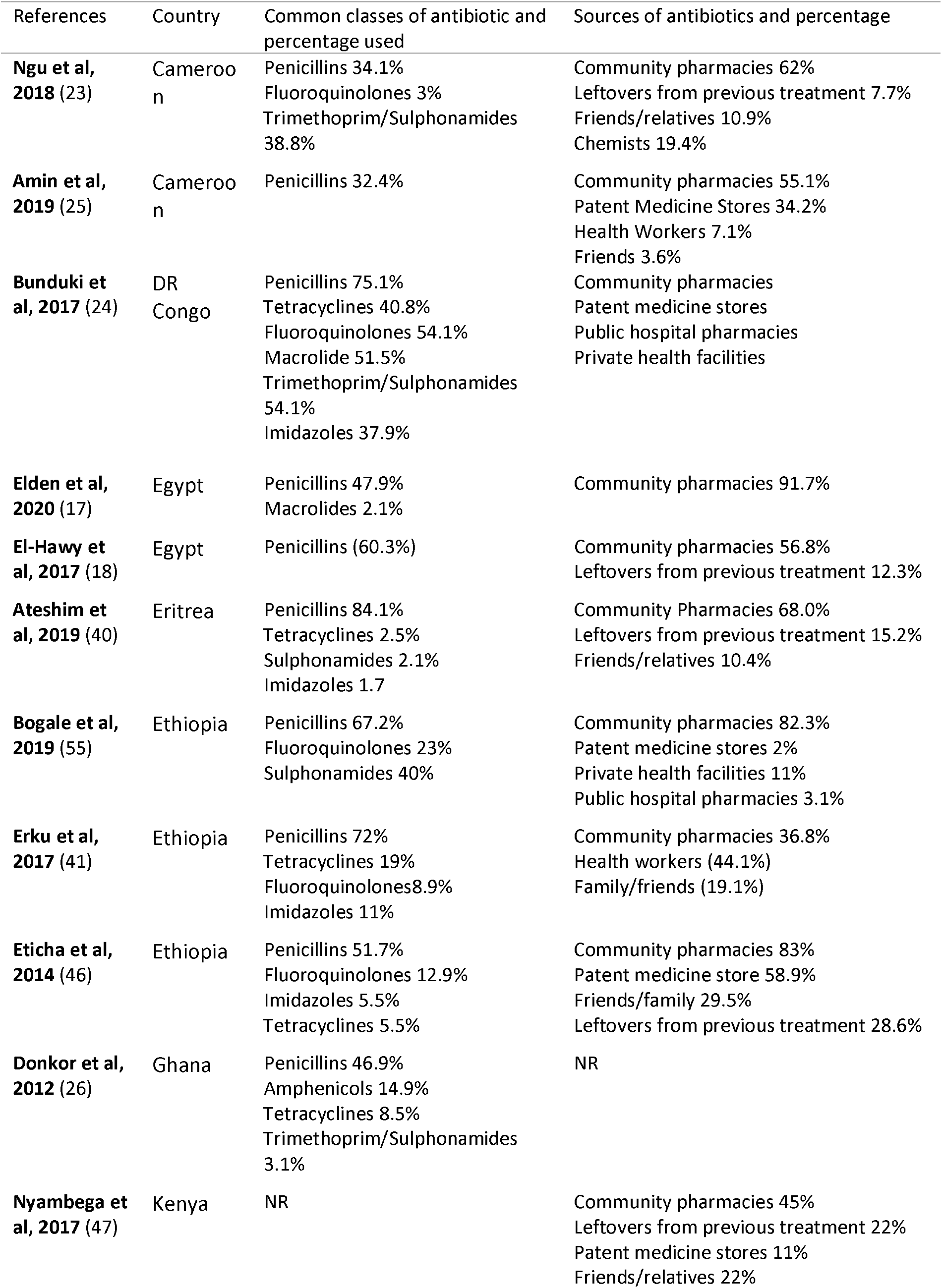
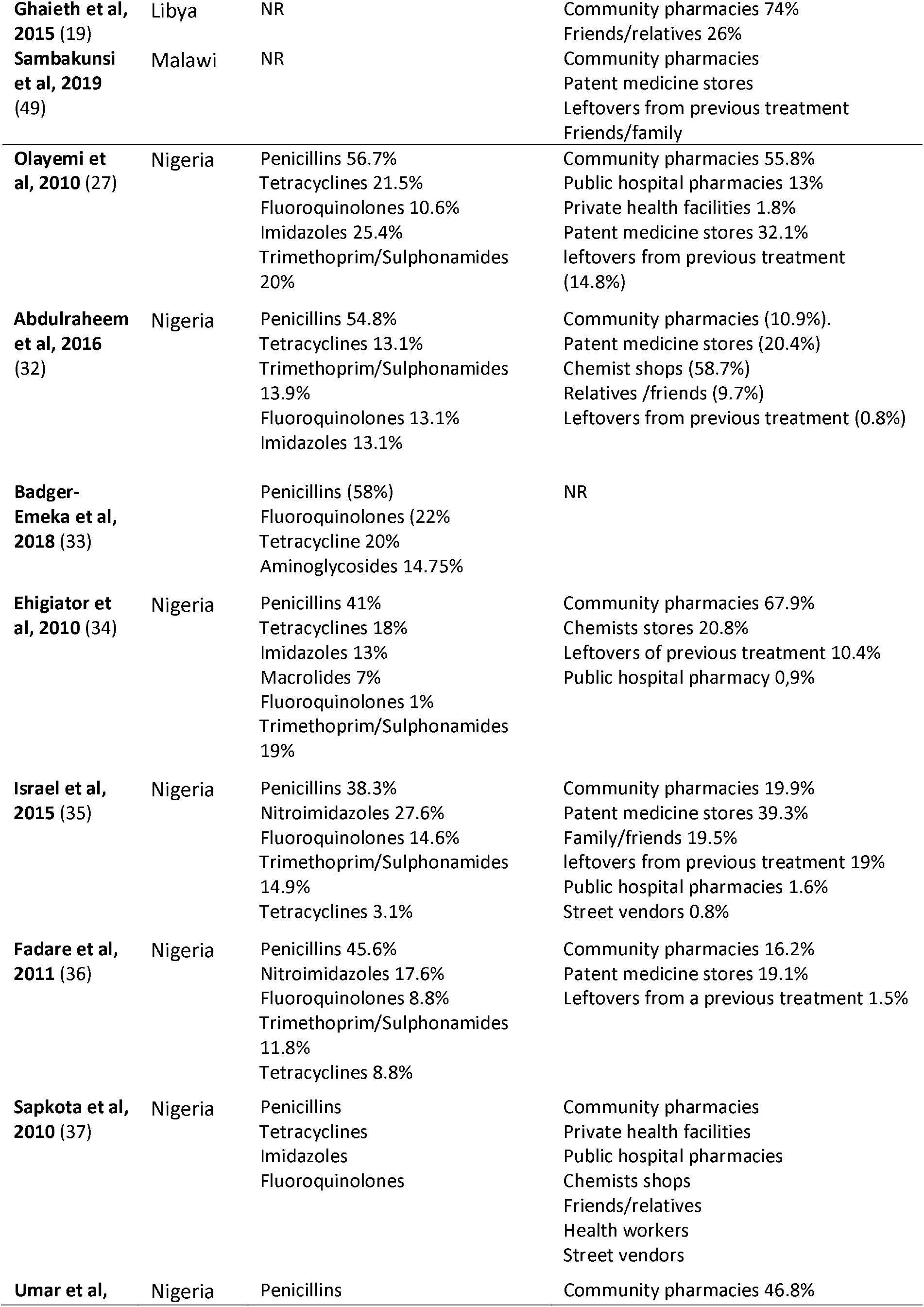
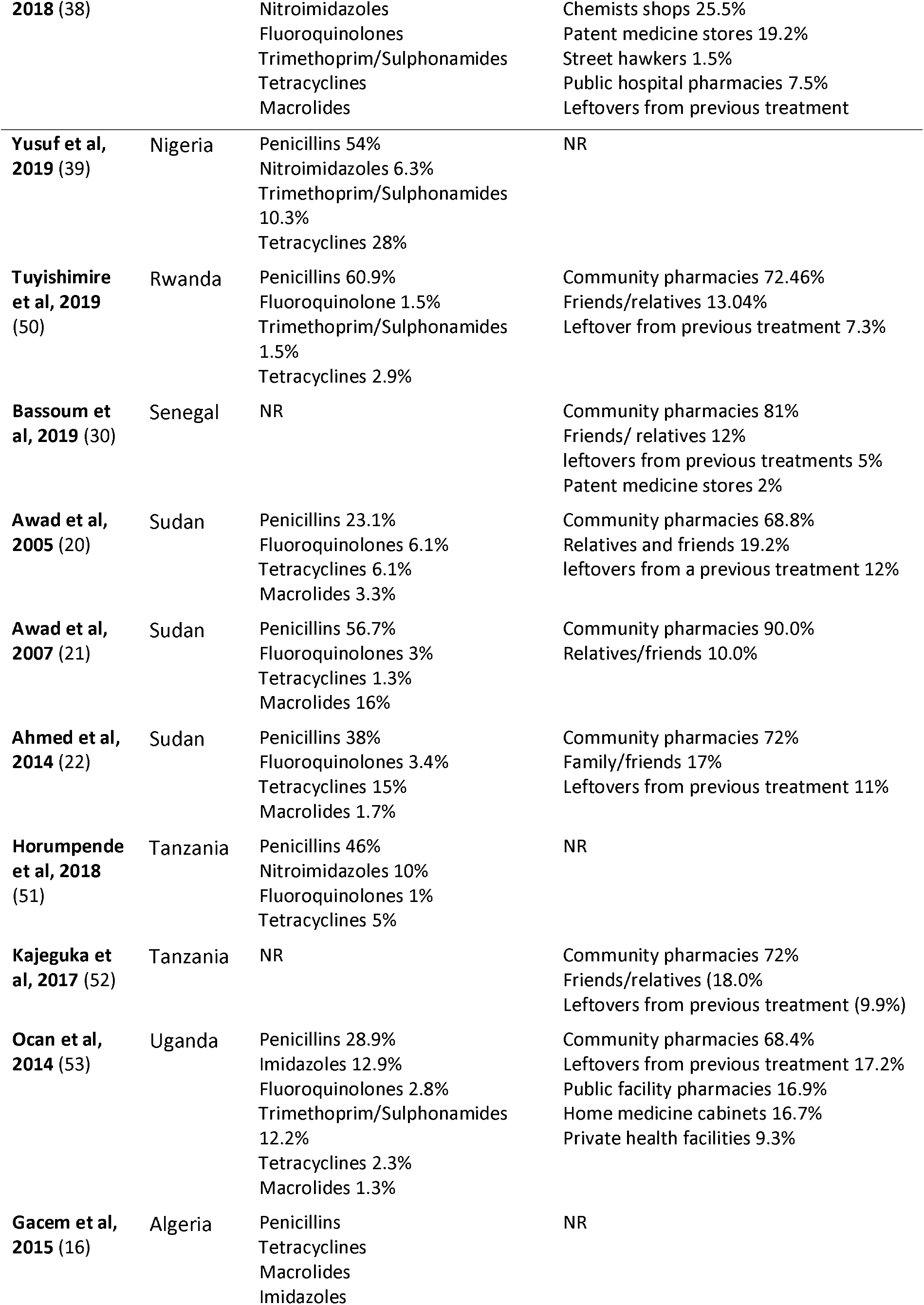
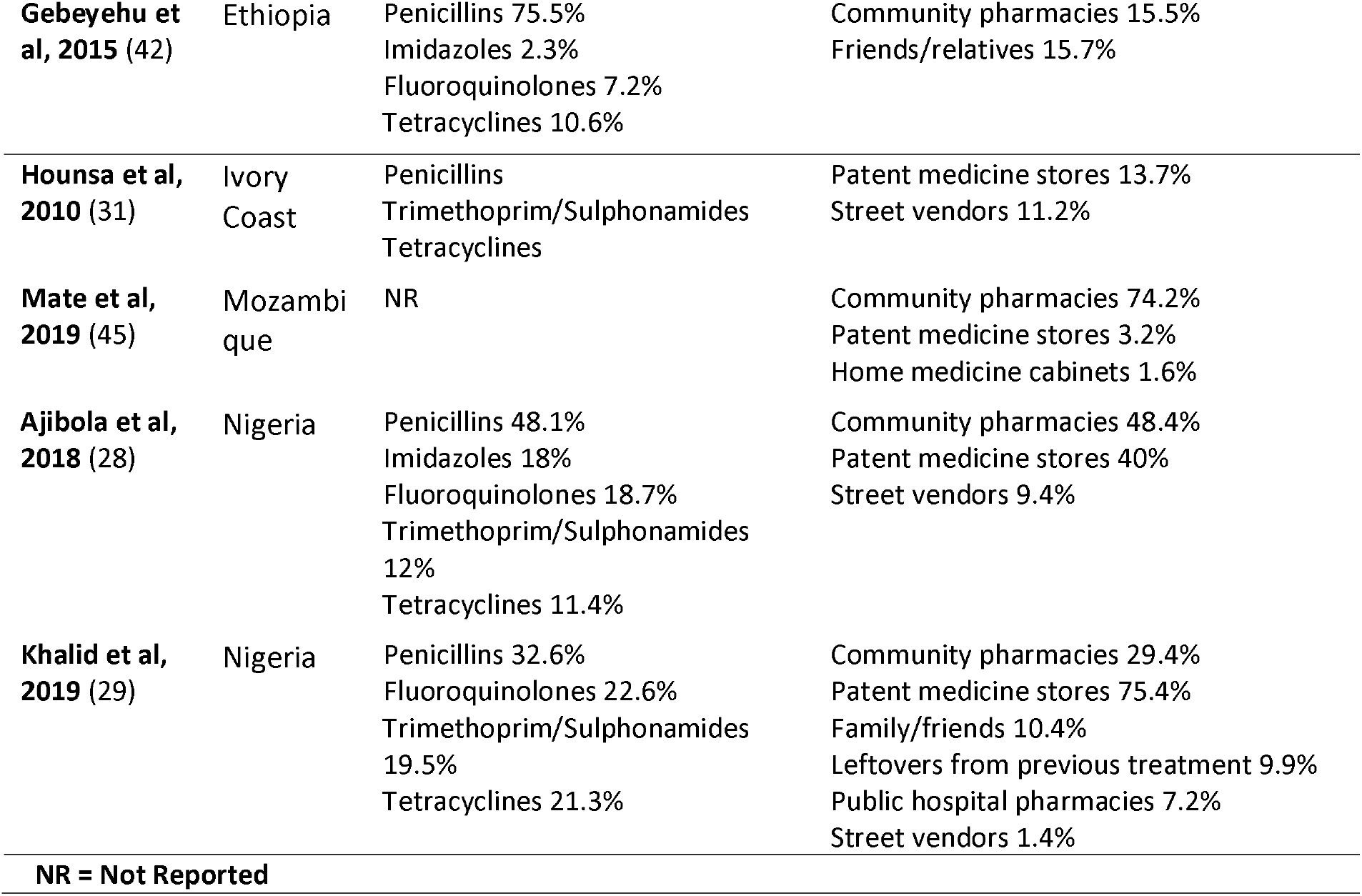
Common antibiotic classes used in self-medication and their sources

**Table 4:**
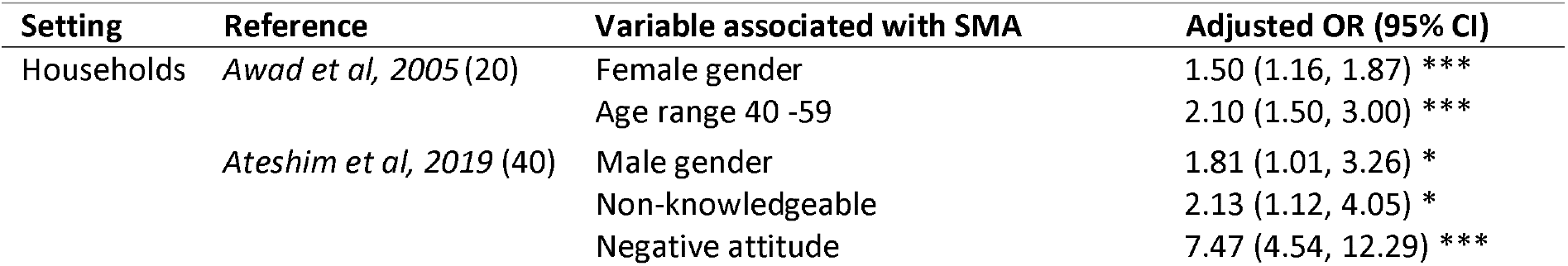
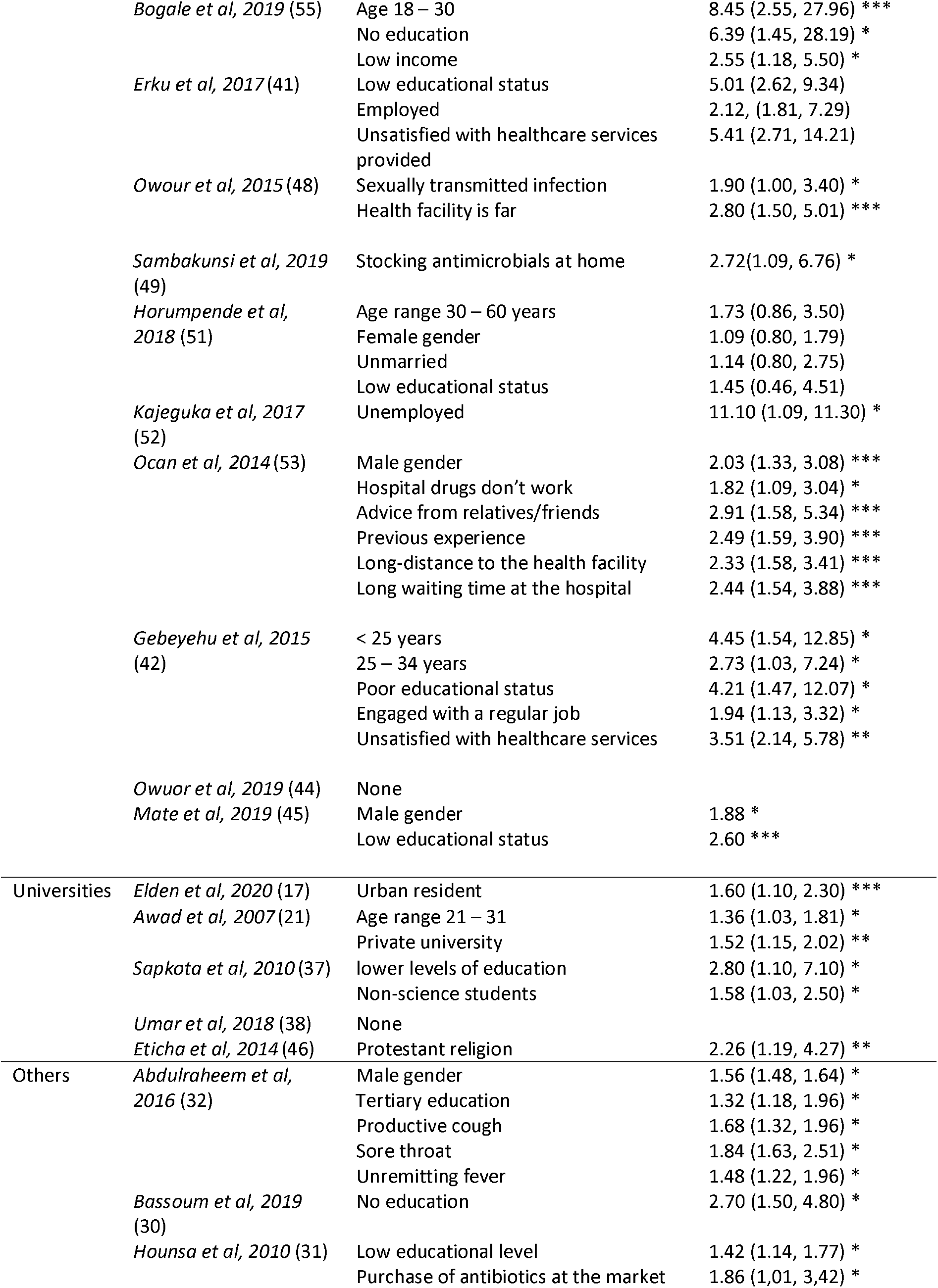
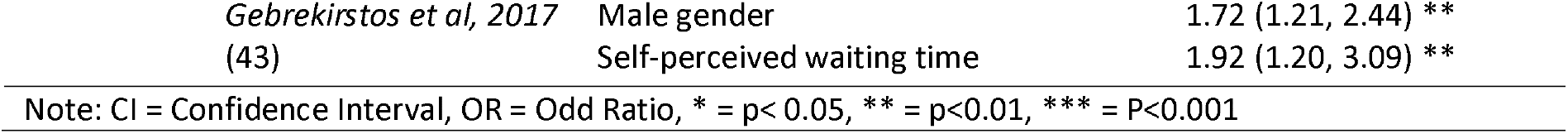
Factors associated with SMA

| Setting | Reference | Variable associated with SMA | Adjusted OR (95% CI) |
| --- | --- | --- | --- |
| Households | *Awad et al, 2005* (20) | Female gender | 1.50 (1.16, 1.87) *** |
|  |  | Age range 40 -59 | 2.10 (1.50, 3.00) *** |
|  | *Ateshim et al, 2019* (40) | Male gender | 1.81 (1.01, 3.26) * |
|  |  | Non-knowledgeable | 2.13 (1.12, 4.05) * |
|  |  | Negative attitude | 7.47 (4.54, 12.29) *** |
|  | *Bogale et al, 2019* (55) | Age 18 – 30 | 8.45 (2.55, 27.96) *** |
|  |  | No education | 6.39 (1.45, 28.19) * |
|  |  | Low income | 2.55 (1.18, 5.50) * |
|  | *Erku et al, 2017* (41) | Low educational status | 5.01 (2.62, 9.34) |
|  |  | Employed | 2.12, (1.81, 7.29) |
|  |  | Unsatisfied with healthcare services provided | 5.41 (2.71, 14.21) |
|  | *Owour et al, 2015* (48) | Sexually transmitted infection | 1.90 (1.00, 3.40) * |
|  |  | Health facility is far | 2.80 (1.50, 5.01) *** |
|  | *Sambakunsi et al, 2019* (49) | Stocking antimicrobials at home | 2.72(1.09, 6.76) * |
|  | *Horumpende et al, 2018* (51) | Age range 30 – 60 years | 1.73 (0.86, 3.50) |
|  |  | Female gender | 1.09 (0.80, 1.79) |
|  |  | Unmarried | 1.14 (0.80, 2.75) |
|  |  | Low educational status | 1.45 (0.46, 4.51) |
|  | *Kajeguka et al, 2017* (52) | Unemployed | 11.10 (1.09, 11.30) * |
|  | *Ocan et al, 2014* (53) | Male gender | 2.03 (1.33, 3.08) *** |
|  |  | Hospital drugs don't work | 1.82 (1.09, 3.04) * |
|  |  | Advice from relatives/friends | 2.91 (1.58, 5.34) *** |
|  |  | Previous experience | 2.49 (1.59, 3.90) *** |
|  |  | Long-distance to the health facility | 2.33 (1.58, 3.41) *** |
|  |  | Long waiting time at the hospital | 2.44 (1.54, 3.88) *** |
|  | *Gebeyehu et al, 2015* (42) | < 25 years | 4.45 (1.54, 12.85) * |
|  |  | 25 – 34 years | 2.73 (1.03, 7.24) * |
|  |  | Poor educational status | 4.21 (1.47, 12.07) * |
|  |  | Engaged with a regular job | 1.94 (1.13, 3.32) * |
|  |  | Unsatisfied with healthcare services | 3.51 (2.14, 5.78) ** |
|  | *Owuor et al, 2019* (44) | None |  |
|  | *Mate et al, 2019* (45) | Male gender | 1.88 * |
|  |  | Low educational status | 2.60 *** |
| Universities | *Elden et al, 2020* (17) | Urban resident | 1.60 (1.10, 2.30) *** |
|  | *Awad et al, 2007* (21) | Age range 21 – 31 | 1.36 (1.03, 1.81) * |
|  |  | Private university | 1.52 (1.15, 2.02) ** |
|  | *Sapkota et al, 2010* (37) | lower levels of education | 2.80 (1.10, 7.10) * |
|  |  | Non-science students | 1.58 (1.03, 2.50) * |
|  | *Umar et al, 2018* (38) | None |  |
|  | *Eticha et al, 2014* (46) | Protestant religion | 2.26 (1.19, 4.27) ** |
| Others | *Abdulraheem et al, 2016* (32) | Male gender | 1.56 (1.48, 1.64) * |
|  |  | Tertiary education | 1.32 (1.18, 1.96) * |
|  |  | Productive cough | 1.68 (1.32, 1.96) * |
|  |  | Sore throat | 1.84 (1.63, 2.51) * |
|  |  | Unremitting fever | 1.48 (1.22, 1.96) * |
|  | *Bassoum et al, 2019* (30) | No education | 2.70 (1.50, 4.80) * |
|  | *Hounsa et al, 2010* (31) | Low educational level | 1.42 (1.14, 1.77) * |
|  |  | Purchase of antibiotics at the market | 1.86 (1.01, 3.42) * |

|  |  |  |
| --- | --- | --- |
| *Gebrekiros et al, 2017* | Male gender | 1.72 (1.21, 2.44) ** |
| (43) | Self-perceived waiting time | 1.92 (1.20, 3.09) ** |

Note: CI = Confidence Interval, OR = Odd Ratio, \* = $p < 0.05$ , \*\* = $p < 0.01$ , \*\*\* = $P < 0.001$

## DISCUSSION

This systematic review analyzed the extent to which SMA is practiced across Africa, the sources of SMA and the main reasons reported why people self-medicate with antibiotics. The prevalence of SMA ranged from 12.1% to 93.9%, whereby the overall median prevalence in Africa is higher than that obtained in South East Asia and the Middle East (3,56). Comparing median prevalence estimates by sub-regions showed that Western Africa reported the highest median prevalence, followed by Northern Africa and Eastern Africa. Previously, the high prevalence of SMA is Africa has been linked to population growth, health inequities and weak healthcare systems coupled with poorly regulated procurement, dispensing and use of antibiotics, and the huge role of an informal health sector (57,58).

Socio-economic determinants of health vary from sub-region to sub-region, and country to country. These are associated with the structure and conditions of health systems, and the health-seeking behaviors of people (57). The higher prevalence estimates that are observed in Africa could be related to the burden of infectious diseases, which is higher than that of HIC warranting a greater use of antibiotics. There is also variation in the way antibiotic sales are regulated across regions and countries. In most African countries, antibiotics are available over-the-counter, and can be obtained from CPs and PMS without prescription, similar to conditions in other LMIC in the Middle East and South East Asia Regions. Poor regulation of antibiotic sales resulting from the absence of policies or laxity in law enforcement makes antibiotics easily available for self-medication (9). A different scenario occurs in HICs, where non-prescription sales of antibiotics is often prohibited with a relatively lower prevalence of SMA observed (3). Law enforcement activities could include government inspections, retention of medical prescriptions in pharmacies, the involvement of pharmacists in designing interventions, and educational interventions (59). Without government inspections, many CPs may tend to dispense antibiotics based on financial motivation and not strictly on medical indications (9).

This review found that CPs and PMS were the main sources of antibiotics used for SMA. Controlling over-the-counter sales of antibiotics in Africa can be a useful strategy to mitigate SMA. This process has proven successful in HIC where that is done by engaging pharmacists in the development of interventions, retention of medical prescriptions in pharmacies, regular inspections of pharmacies by the government, and media campaigns in communities (59). Limiting access to over-the-counter antibiotics without improving access to healthcare in general may not be a tangible solution in resource-limited settings like Africa where many communities are experiencing an absence of medical doctors. This problem can be addressed with task-shifting, thereby authorizing pharmacists and state-registered nurses to prescribe and dispense Access Group antibiotics. Patent medicine stores are community retail stores managed often by non-qualified personnel, and are prohibited from existing in many African countries. Unqualified persons involved in sales of antibiotics do not have sufficient knowledge and skills to properly counsel patients on antibiotics use, control the dosages dispensed, and quality of antibiotics sold. Using leftover antibiotics and old prescriptions is an indication of inappropriate antibiotic usage and a lack of proper education. Preventing reuse of leftovers can be another effective way of preventing SMA (60). This can be achieved by counselling patients when dispensing antibiotics and ensuring that the quantity dispensed corresponds to that prescribed and encouraging the return of uncompleted antibiotics in CPs against financial re-imbursement. Strengthening regulations on dispensing practices and physicians’ sensitization of patients during consultations and physicians will limit this practice and slowly instigate a behavioral change in many people.

Financial constraints, limited access to healthcare and easy access to antibiotics from CPs due to lack of regulatory measures were the most frequently cited motives for SMA in Africa. We identified a nonchalant attitude of healthcare workers towards patients, not involving them in decision making as one of the main causes leading to a lack of trust in healthcare workers. Patient-centered care, though an important component of the acceptability of health services, is still grossly lacking among many health workers especially in Africa (61). Due to the huge patient load, many medical doctors don’t have enough time to properly communicate with patients or caregivers, and they often focus mainly on the biomedical aspects of health and fail to integrate psychosocial aspects of care. Coupled with all the other bottlenecks encountered in the entire healthcare delivery in resource-limited settings, most patients leave the consultation office unsatisfied and this reduces trust and acceptability of health services (61). Many countries in Africa have weak and poorly developed local health systems characterized by lack of facilities and poor quality of service delivery. This negatively affects health-care seeking behaviors and causes many people to go for the option of purchasing antibiotics directly from CPs and PMS which are easy to access and at a reduced cost. Another major reason cited amongst top enablers of SMA is the reliance on past experiences. When people suffer from recurrent or chronic medical problems, they easily develop a habit of self-medicating facilitated in the context where they can get drugs over-the-counter. They often rely on their prior successes, hoping that the outcome will always remain the same with all disease episodes.

SMA in Africa occurs for many different indications and with different antibiotics. Penicillins were the most widely used class of antibiotics used for SMA in Africa. These finding were similar to those reported in other studies (62–64). Penicillins are widely used for SMA because they have lesser side effects and are cheaper to purchase compared to other classes of antibiotics. Even though WHO recommends that Watch Group antibiotics like Fluoroquinolones or Macrolides should be restricted to prescription-only, due to the potential of resistance developing, they were reported as SMA in Africa by many studies. Reserve Group antibiotics were little reported as SMA, presumably because their rare availability and often available as intravenous injections only and has a high cost.

The most common factors associated with SMA were young age, male gender, low education status, and low-income level. Most of these factors are in line with those reported by Ocan and colleagues in a review on developing countries (65). Accessibility, affordability, and conditions of health facilities, as well as the health-seeking behaviors were among the factors identified by Torres and associates in LMIC (17). Low educational status is the most frequently reported factor associated with SMA, warranting the need to promote literacy among communities in Africa as a vital strategy to also reduce SMA. Illiteracy is a driver to the practice of SMA as individuals and entire communities can be less aware of the health risks associated with SMA. A multicenter study carried out in Europe also revealed higher gross domestic product and dispensing the exact quantities of prescribed doses were independently associated with a lower likelihood of SMA, while the perceived availability of antibiotics over-the-counter was the key enabling factor for SMA (60). SMA rates tend to be higher in countries with a lower gross domestic product like African countries. HIC have well-structured health systems with good healthcare infrastructures, adequate access to healthcare services and good health insurance coverage reflecting the high gross domestic product.

## LIMITATIONS

Limitations of this review include unequal distribution and territorial coverage of included studies. All the studies included in this review came from 19 countries out of the 54 countries in Africa with over 80% of the studies from Western Africa and Eastern Africa, while 3 countries provide 50% of the studies. This could explain the relatively high prevalence in Western Africa compared to other sub-regions. However, studies are often carried out when a problem is suspected. There are some limitations introduced by the potential biases from individual studies. Fifteen out of the 40 studies included had a moderate risk of bias. These biases resulted from variations in the method of analysis and recall period, selection and social desirability, scientific rigor, study population, and sample size. Other issues that lead to bias were the use of non-random sampling procedures to recruit participants, and failure to validate the survey questionnaires in some studies. Also, some studies did not meet the correct case definition of SMA, as they indiscriminately used either antimicrobials and antibiotics and without specifying the duration. Many studies had a recall period used during data collection of more than 6 months, while some studies did not report the recall period used in the study at all.

## CONCLUSION

The prevalence of SMA in Africa is high and varies across sub-regions with Western Africa having the highest prevalence. Drivers for SMA comprise of socio-economic factors elucidated by low educational status and financial constraint, limited access to healthcare characterized by high out-of-pockets payments, absence of patient-centered care, and inadequate policies regulating the sales of antibiotics or poor implementation of existing regulations. There will be no one-size-fits-all strategy to address SMA in Africa ensuring effective and sustainable control. Tackling this problem therefore requires a multifaceted approach that is user-centered and context-specific, addressing various actors and stakeholders ranging from antibiotic users to dispensers and policy makers.

## Data Availability

This is a systematic review

## LIST OF ABBREVIATIONS

SMA: Self Medication with antibiotics
CP: Community Pharmacies
PMS: Patent Medicines Stores
LMIC: Low and Middle Income Countries
HIC: High-income Countries

## DECLARATIONS

### Ethics approval and consent to participate

Not applicable

## Consent for publication

Not applicable

## Availability of data and materials

Not applicable

## Authors’ contributions

EVY carried out the review of articles, analysed and interpreted data, and wrote the original manuscript. BI did arbitration of the review process, interpreted data and contributed in writing the original manuscript. JNF acted as the second arbitrary of the review process. BL and MS were major contributors in writing the final manuscript. All authors read and approved the final manuscript.

## Acknowledgements

Authors acknowledge Tine Verdonck for her contribution in analysing and interpretation of meta-analysis.

